# Immune imprinting and vaccination interval underly XBB.1.5 monovalent vaccine immunogenicity

**DOI:** 10.1101/2025.02.09.25321965

**Authors:** Xammy H. Wrynla, Timothy A. Bates, Mila Trank-Greene, Mastura Wahedi, Audrey Hinchliff, Marcel E. Curlin, Fikadu G. Tafesse

## Abstract

As COVID-19 transitions into endemicity and vaccines are annually updated to circulating SARS-CoV-2 lineages such as JN.1, exposure intervals and immune imprinting become critical considerations for vaccination strategy. Imprinting by the ancestral spike protein has been observed with the bivalent Wuhan-Hu-1/BA.4-5 vaccine and its persistence can be further evaluated in the context of the more recent XBB.1.5 monovalent vaccine. We assessed antibody responses in individuals who received three to four doses of Wuhan-Hu-1, one dose of bivalent Wuhan-Hu-1/BA.4-5, and one dose of XBB.1.5 vaccine (bivalent recipients). We compared these to individuals who received three to four doses of Wuhan-Hu-1 and one dose of XBB.1.5 vaccine without prior bivalent vaccination (bivalent non-recipients). Before XBB.1.5 vaccination, bivalent non-recipients demonstrated decreased breadth and potency of neutralizing antibodies compared to recipients, but at post-vaccination exhibited greater boosting of neutralizing antibodies against XBB.1.5 (18.4X versus 6.2X), EG.5.1 (30.9X versus 7.0X), and JN.1 (9.2X versus 3.7X) variants with trends toward higher neutralizing titers and comparable variant cross-neutralization. Increased boosting in non-recipients were similarly observed for IgA and total IgG/A/M isotypes binding the spike receptor-binding domain but not IgG nor IgM. Bivalent non-recipients had longer intervals between exposures, which has been reported to enhance antibody boosting; however, bivalent receipt and interval were tightly linked variables, preventing the isolation of individual contributions to boosting. Nonetheless, significant “back-boosting” of ancestral SARS-CoV-2 titers upon XBB.1.5 vaccination in both participant groups indicate that immune imprinting continues to affect contemporary vaccines. Altogether, our findings highlight imprinting and exposure intervals as important phenomena underlying variant-adapted COVID-19 vaccine immunogenicity.

## INTRODUCTION

The World Health Organization (WHO) declared the end of COVID-19 as a public health emergency in May 2023 (*1*), but SARS-CoV-2 variants continue to emerge and spread (*2*). To maintain population immunity, the XBB.1.5 monovalent vaccine was subsequently approved in 2023 and was shown to successfully induce cellular responses and neutralizing antibodies against circulating variants (*3–6*). The JN.1-adapted vaccines were further formulated to match prevalent strains in 2024 (*7*). However, the diversity and potency of new immune responses generated through updated COVID-19 vaccines may be influenced by immune imprinting following exposure to historic spike proteins during prior vaccination or natural infection (*8*).

Immune imprinting has been extensively explored for influenza and is characterized by reduced induction of neutralizing antibodies against novel variants of a viral antigen upon exposure. “Back-boosting” is also highly characteristic of imprinting and describes boosting of titers against ancestral antigen upon exposure to a novel variant (*8*). Recognition of immune imprinting in the context of COVID-19 initially arose with observations that infection resulted in “back-boosting,” of binding antibodies against seasonal coronaviruses OC43 and HKU1 (*9*). Ancestral Wuhan-Hu-1 infection prior to Omicron BA.1 infection resulted in poorer Omicron-targeting antibody responses compared to infection with BA.1 alone (*10*). Similarly, vaccination with monovalent Wuhan-Hu-1 prior to infection with Alpha or Delta variants generated weaker variant-specific responses than in unvaccinated, infected individuals (*11*). As the first widely administered variant-adapted vaccine, the bivalent Wuhan-Hu-1/BA.4-5 vaccine failed to significantly enhance Omicron-neutralizing antibodies compared to monovalent Wuhan-Hu-1 boosting, likely due to imprinting by the ancestral spike protein (*12–14*). Consistent with this observation, minimal additional protection was provided against COVID-19 infection and hospitalization in bivalent versus Wuhan-Hu-1 monovalent recipients (*15*). More recently, studies have demonstrated that targeting immune responses to ancestral SARS-CoV-2 by means of additional booster doses increases susceptibility to reinfection with emerging Omicron variants (*16*). Though these reinfection study designs may be confounded by selection bias (*17*), reports assessing XBB.1.5 vaccine-induced pseudovirus neutralizing antibodies indeed observe the “back-boosting” of responses against historic variants that is characteristic of immune imprinting (*5*, *8*).

To further evaluate the impact of immune imprinting on XBB.1.5 vaccine immunogenicity, we identified a group of individuals who did not previously receive the bivalent vaccine in 2022 but did receive the XBB.1.5 monovalent vaccine in 2023 (bivalent non-recipients). Between bivalent non-recipients and participants who received the bivalent vaccine prior to XBB.1.5 vaccination (bivalent recipients), we compared serum neutralizing antibody titers that effectively block cellular infection by live clinical isolates of SARS-CoV-2 representing ancestral (WA1), vaccine-matched (XBB.1.5), and emergent variants (EG.5.1 and JN.1) and measured IgG, IgA, IgM, and total IgG/A/M isotypes targeting the ancestral spike receptor-binding domain (RBD).

## RESULTS

### Cohort and study design

49 participants were initially recruited; however, serum samples were analyzed for anti-nucleocapsid seropositivity by enzyme-linked immunosorbent assay (ELISA) to screen for recent prior infection. Nine seropositive participants were excluded from the final cohort. Within the final cohort of 40 participants (**Table 1**), 30 received three or four doses of monovalent Wuhan-Hu-1 followed by one dose of bivalent Wuhan-Hu-1/BA.4-5 vaccine (bivalent recipients), and 10 received three or four doses of monovalent Wuhan-Hu-1 vaccine (bivalent non-recipients) (**Figure 1A**). At time of recruitment, all participants received the XBB.1.5 monovalent mRNA vaccine manufactured by Moderna Therapeutics. Suspected and rapid antigen-confirmed infection history were collected from participants. Serum samples were collected from each participant prior to XBB.1.5 vaccination and again at an average of 22 days post-vaccination. These were tested for 50% effective antibody IgG, IgA, IgM, and total IgG/A/M isotype concentrations (EC50) by ELISA against the ancestral spike RBD as well as 50% live SARS-CoV-2 neutralizing titers by focus reduction neutralization tests (FRNT50) against the ancestral WA1, vaccine-matched XBB.1.5, and emergent EG.5.1 and JN.1 strains.

**Table 1.** Final Cohort Demographics.

| **Group** | **Bivalent Recipients** | **Bivalent Non-recipients** |
| --- | --- | --- |
| **# Participants** | 30 | 10 |
| **# Female (%)** | 21 (70%) | 6 (60%) |
| **# Male (%)** | 9 (30%) | 4 (40%) |
| **Mean age (range)** | 52.2 (26-74) | 50.8 (35-71) |
| **Mean days between serum collection (range)** | 22.1 (14-38) | 20 (14-41) |
| **Mean interval since prior vaccination (range)** | 361.2 (154-405) | 706.9 (555-742) |
| **Prior vaccine type** | bivalent Wuhan-Hu-1/BA.4-5 | monovalent Wuhan-Hu-1 |
| **Mean # of prior infections (range)** | 0.8 (0-3) | 0.9 (0-2) |
| **Median # of prior infections (range)** | 1 (0-3) | 1 (0-2) |

**Fig. 1.**
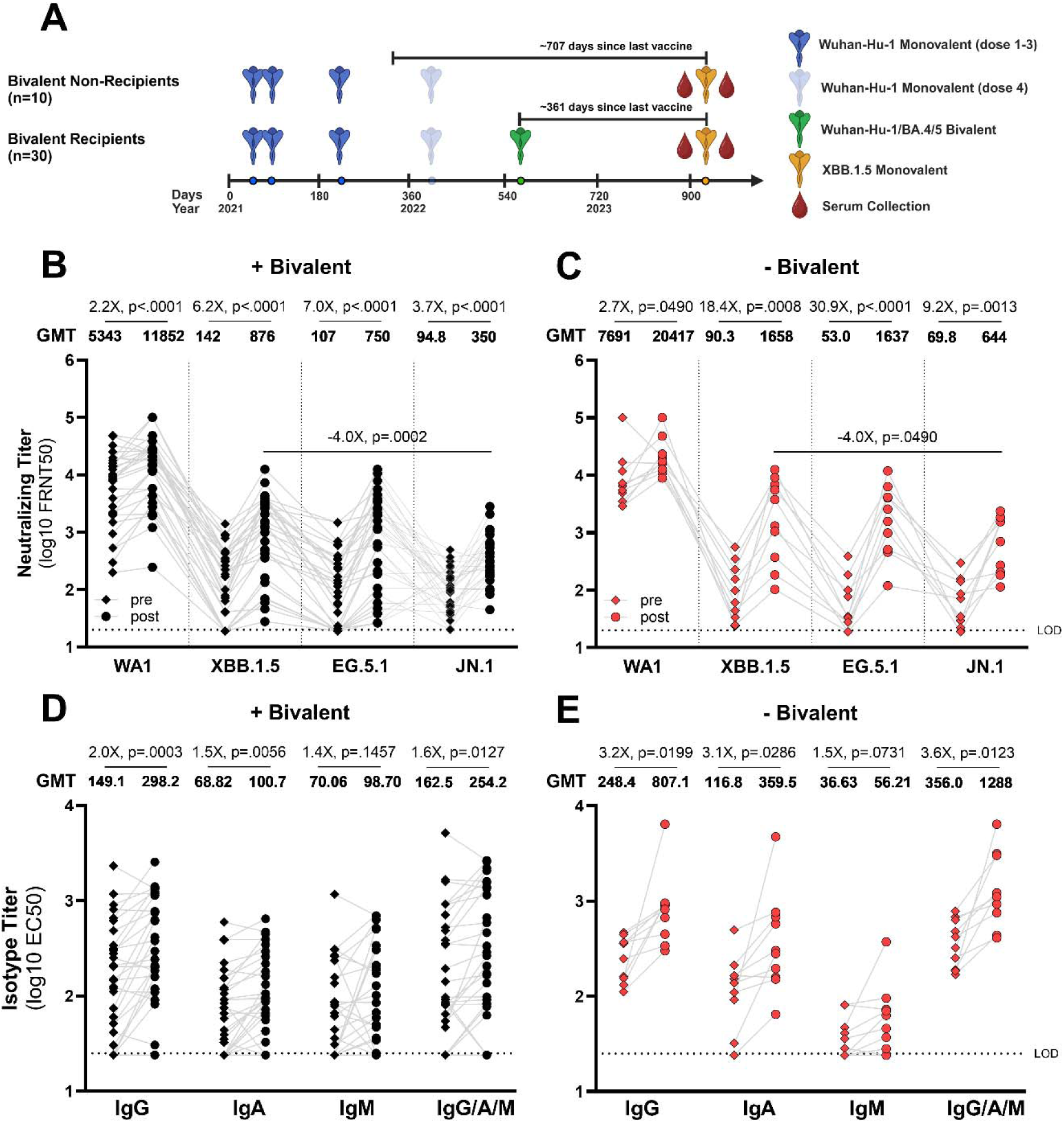
XBB.1.5 monovalent vaccination boosts antibody responses. (**A**) Pre- and post-XBB.1.5 vaccination sera were collected from healthcare workers who previously received the bivalent vaccine or not. Live SARS-CoV-2 neutralization by serum antibodies was assessed by focus reduction neutralization test (FRNT) and reported as FRNT50s for bivalent recipients (**B**) and non-recipients (**C**). Serum antibody isotype titers against ancestral spike RBD were determined by enzyme-linked immunosorbent assay (ELISA) and reported as EC50 for bivalent recipients (**D**) and non-recipients (**E**). The dotted lines indicate assay lower limits of detection. Geometric mean titers (GMT) for each bar were calculated in GraphPad Prism. Fold changes were calculated by dividing the post-XBB.1.5 vaccination titer by pre-vaccination titer. Reported p-values are the result of a one-way repeated measures ANOVA (B, C) or restricted maximum likelihood model (D, E) with Holm-Sidak multiple comparisons correction.

### XBB.1.5 monovalent vaccination boosts antibody titers

In both bivalent recipient and non-recipient groups, the XBB.1.5 monovalent vaccine successfully boosted neutralizing titers against XBB.1.5 (recipient p<.0001, non-recipient p=.0008), EG.5.1 (recipient p<.0001, non-recipient p<.0001), and JN.1 (recipient p<.0001, non-recipient p=.0013) and “back-boosted” titers against the ancestral WA1 strain (recipient p<.0001, non-recipient p=.0490). Of note, the JN.1 variant, from which currently dominant strains emerged, demonstrated significant escape from vaccinated sera with neutralizing titers that were 4-fold lower than against the vaccine-matched XBB.1.5, regardless of vaccine history (recipient p=0.0002, non-recipient p=0.0490) (**Figure 1B, 1C**). In both groups, the XBB.1.5 vaccine similarly “back-boosted” IgG (recipient p=.0003, non-recipient p=.0199), IgA (recipient p=.0056, non-recipient p=.0286), and total IgG/A/M (recipient p=.0127, non-recipient p=.0123) isotype antibodies targeting the ancestral spike RBD, but did not significantly boost RBD-specific IgM (recipient p=.1457, non-recipient p=.0731) (**Figure 1D, 1E**).

### Bivalent non-recipients exhibit diminished neutralizing breadth and potency prior to XBB.1.5 vaccination

To visually assess breadth of neutralizing antibody responses, neutralizing titers against the XBB.1.5, EG.5.1, and JN.1 variants were plotted against ancestral WA1 titers. 95% confidence ellipses were drawn to delineate the distribution of each participant group. Antibodies in both groups demonstrated a strong preference toward neutralizing WA1 over variants at pre-vaccination, but these cross-neutralizing responses were improved by XBB.1.5 vaccination as denoted by the upward shift toward increased variant-specific titers **(Figure 2A, 2B, 2C, 2D, 2E, 2F**).

**Fig. 2.**
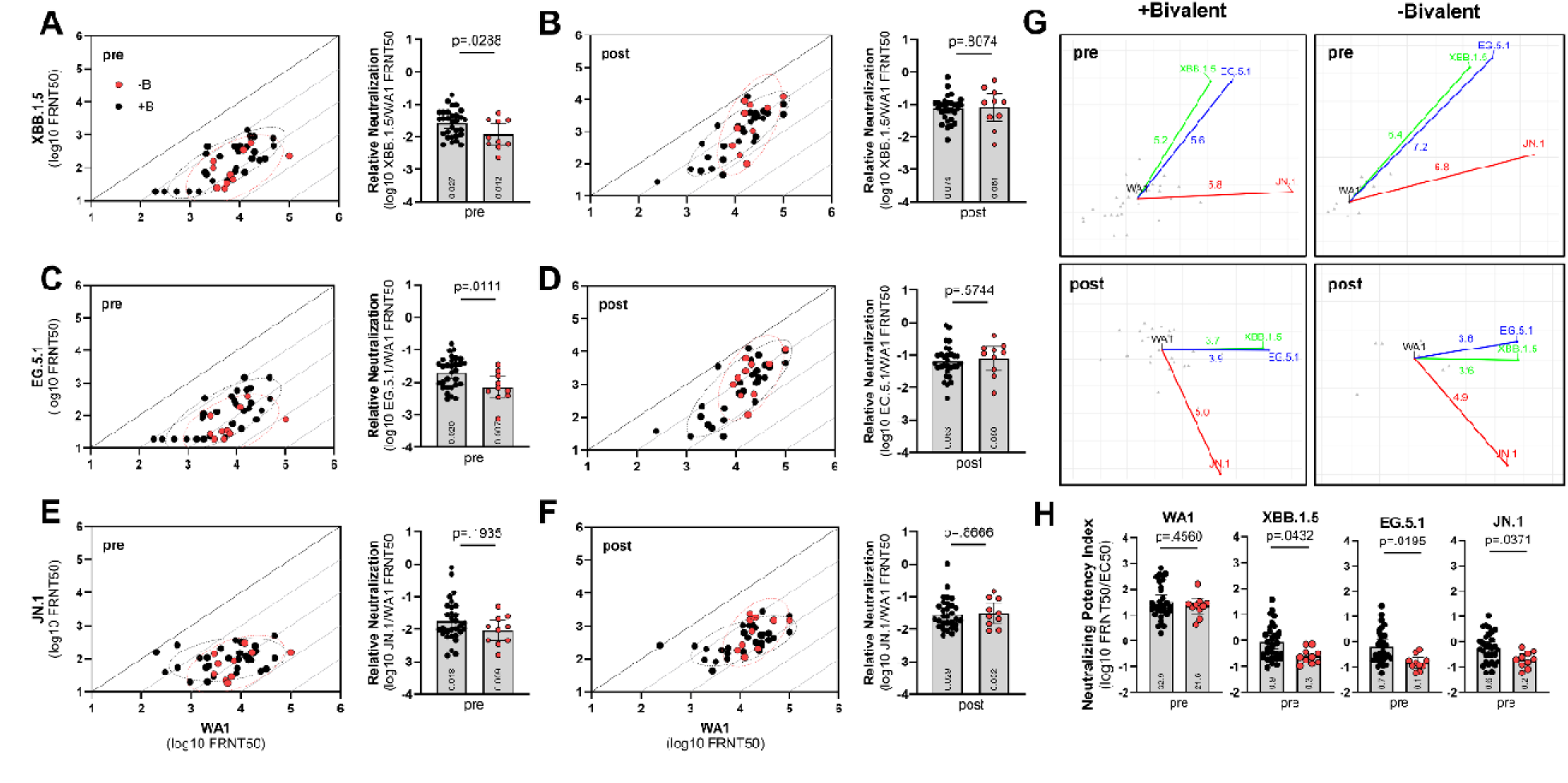
Bivalent non-recipients exhibit diminished variant cross-neutralization and neutralizing potency at pre-vaccination. For each participant, pre- and post-vaccination neutralizing titers against live, clinical isolates of XBB.1.5 (**B, E**), EG.5.1 (**C, F**), or JN.1 (**D, G**) variants were plotted against neutralizing titers against WA1. The black dotted line represents equal neutralization, and the dotted ellipses represent 95% confidence ellipses. Relative neutralization ratios were also calculated by dividing neutralizing titers against each variant by those against WA1. (**H**) Antigenic cartographs were generated using the Racmacs packages in R v4.3.1 for bivalent recipients and non-recipients at each timepoint. Arrows and corresponding values represent quantified map distances between WA1 and variants. Each square-length represents ∼2-fold changes in FRNT50 neutralizing titer. Gray triangles represent positions for each serum sample. (**I**) Neutralizing potency index (NPI) for pre-XBB.1.5 vaccination (pre) serum samples were calculated by dividing the FRNT50 against WA1, XBB.1.5, EG.5.1, or JN.1 by total IgG/A/M EC50 for each participant. For all bar plots, the plotted geometric mean values are displayed. Error bars are geometric means with 95% confidence intervals. Reported p-values are the result of unpaired t tests.

In comparing groups at the pre-vaccination timepoint, bivalent non-recipients had a slight skew toward WA1 neutralization relative to recipients. To quantitate these visual observations, relative neutralization ratios were calculated by dividing the neutralizing titer against a given variant by WA1 titers. At pre-vaccination, bivalent non-recipients had significantly lower relative neutralization ratios for XBB.1.5 (p=.0305) and EG.5.1 (p=.0111) compared to recipients, possibly due to absence of prior vaccine exposure to the BA.4/5 spike variant which would enhance Omicron cross-neutralizing responses. The trend for JN.1 was similar but not statistically significant (p=.1935) (Figure 2A, 2B, 2C). However, after XBB.1.5 vaccination, the bivalent non-recipient group clustered to the upper, right distribution of the total cohort, suggesting a trend toward improved cross-neutralization in addition to overall potency of neutralization. However, this enhancement in cross-neutralization was not significant upon comparing relative neutralization ratios (Figure 2D, 2E, 2F).

Antigenic cartography was performed to further evaluate variant cross-neutralization. Here, neutralizing titer values against each SARS-CoV-2 strain for each participant serum sample were used to generate antigenic maps. One map was generated for each group (bivalent recipient and non-recipient) and at each timepoint (pre- and post-vaccination). One square-length map distance represents a ∼2-fold change in FRNT50 neutralizing titer and is a measure of antigenic difference. Across all maps, XBB.1.5 and EG.5.1 were positioned closest together and appear most antigenically similar from the perspective of serum titers; this is consistent with the fact that EG.5.1 is a closely related descendent of the XBB.1.5 lineage while WA1 and JN.1 are phylogenetically more distant (*18*). In both groups, map distances between WA1 and variants are shorter at post-vaccination compared to pre-vaccination, demonstrating successful boosting of cross-neutralizing responses by the XBB.1.5 monovalent vaccine (**Figure 2G**).

Unsurprisingly, bivalent non-recipients exhibited longer map distances than recipients at pre-vaccination between WA1 and XBB.1.5 (6.4 versus 5.2), EG.5.1 (7.2 versus 5.6), and JN.1 (6.8 versus 5.8), suggesting diminished cross-neutralization from lack of prior vaccination with the BA.4/5 spike variant. At post-vaccination, however, bivalent non-recipients had slightly shorter but overall similar map distances between WA1 and XBB.1.5 (3.6 versus 3.7), EG.5.1 (3.6 versus 3.9), and JN.1 (4.9 versus 5.0) (Figure 2G). These analyses suggest that despite reduced pre-vaccination cross-neutralization due to lack of the bivalent vaccine, non-recipients recover comparable cross-neutralization to bivalent recipients upon XBB.1.5 vaccination.

Finally, neutralizing potency of antibody responses at pre-vaccination was measured by assessing the neutralizing potency index (NPI), which is calculated by dividing the serum neutralizing titer for a given variant by the total IgG/A/M titer. Biologically, each variant-specific NPI quantifies the subset of spike RBD-binding antibodies that lead to neutralization of infection by the assessed variant. The neutralizing potency of serum antibodies were significantly lower against XBB.1.5 (p=.0432), EG.5.1 (p=.0195), and JN.1 (p=.0371) but not WA1 (p=.4560) in bivalent non-recipients prior to XBB.1.5 vaccination. As with significantly diminished variant cross-neutralization, reduced neutralizing potency is likely the result of lacking exposure to the BA.4/5 spike variant and highlights the importance of bivalent vaccination in enhancing Omicron-targeting responses (**Figure 2H**).

### XBB.1.5 vaccine elicits greater boosting of variant-neutralizing antibodies in bivalent non-recipients

Pre-vaccination neutralizing titers against the contemporary variants trended lower in bivalent non-recipients against XBB.1.5 (1.6-fold; p=.3107), EG.5.1 (2-fold; p=.1040), and JN.1 (1.4-fold; p=.3792), consistent with greater waning of antibody responses in this group due to the missing bivalent vaccine and longer time interval since prior vaccination. Strikingly, however, after final vaccination, trends toward higher absolute titers were observed in the bivalent non-recipient group against WA1 (1.7-fold; p=0.1158), XBB.1.5 (1.9-fold; p=0.3188), EG.5.1 (2.2- fold; p=0.1773), JN.1 (1.8-fold; p=0.1500), suggesting an anchoring effect due to immune imprinting in bivalent recipients (Figure 1B, 1C). Although these differences in post-vaccination absolute titers between bivalent recipients and non-recipients did not reach statistical significance, boosting of variant-specific neutralizing antibodies (i.e. fold-change after vaccination) was significantly greater in non-recipients against XBB.1.5 (18.4X versus 6.2X, p=0.0134), EG.5.1 (30.9X versus 7.0X, p=0.0027), and JN.1 (9.2X versus 3.7X, p=0.0389). However, no significant difference was observed against WA1 (2.7X versus 2.2X, p=0.5910) (**Figure 3A, 3B, 3C, 3D**).

**Fig. 3.**
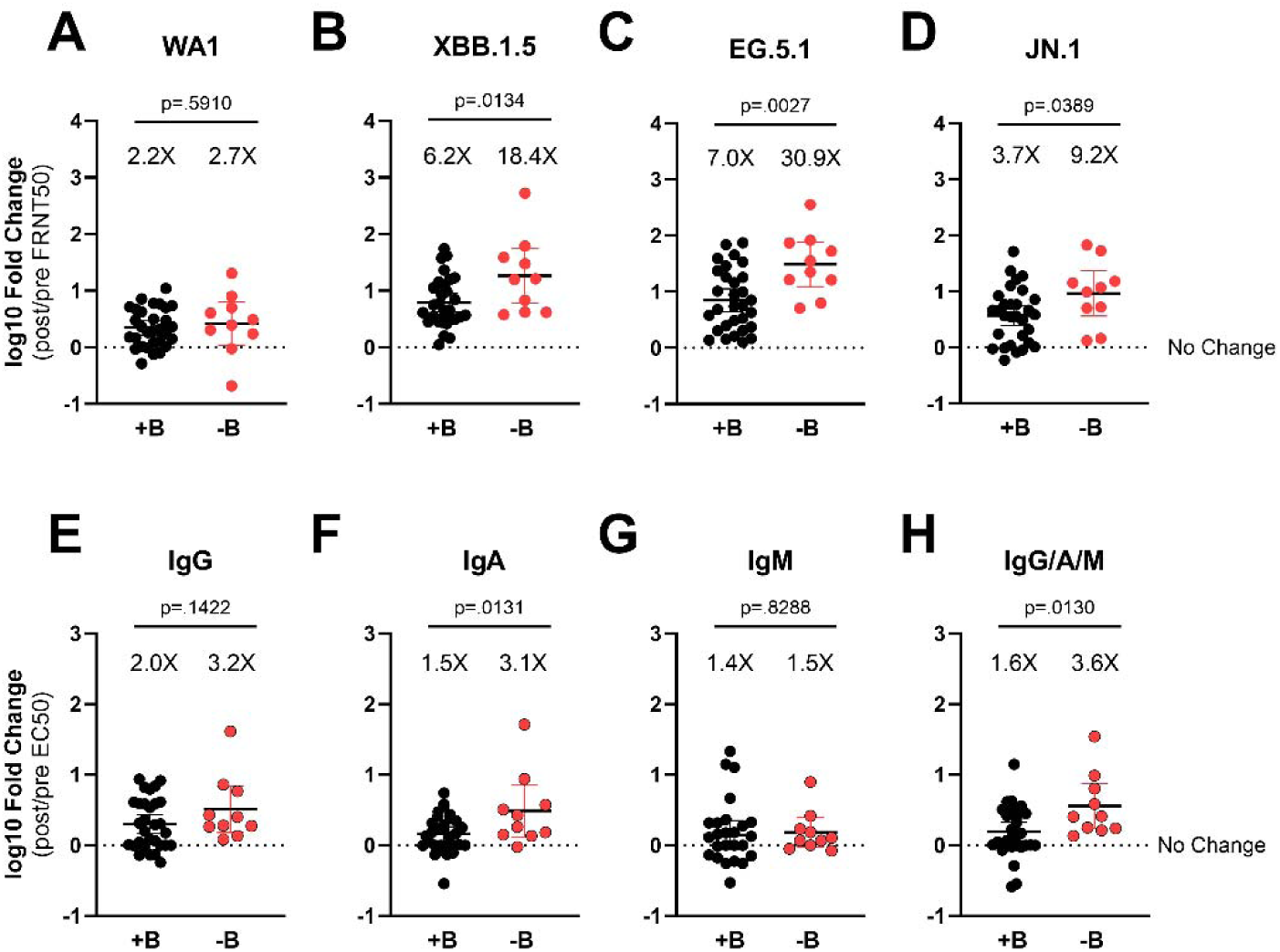
Bivalent non-recipients exhibit stronger boosting of variant-neutralizing and spike RBD-binding isotype titers. Live SARS-CoV-2 neutralizing antibodies were quantified by focus reduction neutralization test (FRNT) and reported as FRNT50s. Fold changes were calculated by dividing the post-XBB.1.5 vaccination titer by pre-vaccination titer for each participant against WA1 (**A**), XBB.1.5 (**B**), EG.5.1 (**C**), and JN.1 (**D**). Ancestral spike RBD-binding antibodies were determined by enzyme-linked immunosorbent assay (ELISA) and reported as EC50s. Fold-changes were calculated for IgG (**E**), IgA (**F**), IgM (**G**), and total IgG/A/M (**H**). The dotted lines indicate no change in titer from pre- to post-vaccination (fold change=1). Error bars show geometric means with 95% confidence intervals. Reported p-values are the result of unpaired t tests.

The bivalent recipient group was further stratified based upon having received either three or four doses of ancestral monovalent vaccine prior to the bivalent vaccine. Using an ANOVA follow-up test called “test for linear trend” (a.k.a. “test for linear contrast,” “post-test for trend”), which statistically assesses systematic decreases in left-to-right column order, these recipient subgroups were then compared to the bivalent non-recipient group in order to assess the effect of ancestral spike vaccinations on XBB.1.5 vaccine immunogenicity. This revealed a generally inverse relationship between number of ancestral spike vaccinations and absolute post-titer that approached significance for XBB.1.5 (p=.0803) and EG.5.1 (p=.0504) while boosting had similar and statistically significant relationships for XBB.1.5 (p=.0124) and EG.5.1 (p=.0030). Within the bivalent recipient group, however, having 3 or 4 doses of ancestral spike resulted in similar responses against JN.1 and thus lack of linear trend in terms of absolute post-titer (p=.1875) and boosting (p=.1219), suggesting that vaccine immunogenicity against JN.1 and strains emerging from this distinct lineage may suffer less from ancestral spike imprinting. While the number of ancestral vaccinations had an impact on absolute post-titers against WA1 (p=0.0323), no difference was observed in boosting (p=.6491), likely due to relatively high baseline neutralizing antibody titers from repetitive vaccine- and infection-mediated “back-boosting” across all participant groups (**Supplemental Figure 1**).

### XBB.1.5 vaccine elicits greater boosting and absolute ancestral spike-binding isotype titers in bivalent non-recipients

Pre-vaccination titers trended higher for bivalent non-recipients despite longer interval since prior vaccination for IgG (1.7-fold; p=.0926), IgA (1.7-fold; p=.1283), and total IgG/A/M (2.2-fold; p=.0236) (Figure 1D, 1E). Our assay was optimized to detect antibody isotypes targeting the ancestral spike RBD and, though unexpected, these higher pre-vaccination titers in the non-recipient group were validated by similarly elevated neutralizing titers against the ancestral WA1 strain (Figure 1B, 1C, Supplemental Figure 1A). These increased pre-vaccination responses against ancestral antigen in bivalent non-recipients may be explained by natural infection histories skewed toward older strains. In other words, non-recipients opted out of the bivalent vaccine and were thus more susceptible to infection by early or pre-Omicron lineages that circulated in 2022. Infection by these lineages would more likely maintain responses against older spike variants, whereas an infection history more dominated by post-XBB Omicron would more drastically draw responses away from ancestral antigen. However, it is important to note that both bivalent recipient and non-recipient groups had similar overall numbers of prior infection.

At post-vaccination, absolute titers were significantly higher in the bivalent non-recipient group for IgG [2.7-fold; p=.0098], IgA [3.6-fold; p=.0089], and total IgG/A/M [5.1-fold; p=.0004] (Figure 1D, 1E). Furthermore, more profound boosting of isotypes was observed in bivalent non-recipients than in recipients for IgA (3.1X versus 1.5X, p=0.0131) and total IgG/A/M (3.5X versus 1.6X, p=0.0130), though weaker and not statistically significant for IgG (3.2X versus 2.0X, p=0.1422) and indifferent for IgM (1.5X versus 1.4X, p=0.8288) (**Figure 3E, 3F, 3G, 3H**).

When stratifying study participants by number of ancestralWuhan-Hu-1 doses received prior to bivalent vaccination and performing ANOVA follow-up column “tests for linear trend,” significant inverse relationships were found with post-vaccination titers for IgG (p=.0222), IgA (p=.0003), and total IgG/A/M (p=.0014) as well as for boosting of IgA (p=.0154). This relationship approached statistical significance for boosting of total IgG/A/M (p=.0799). No relationship was observed for absolute IgM post-titer (p=.4371) nor boosting of IgG (p=.6057) and IgM (p=.9400) (**Supplemental Figure 2**).

### Bivalent receipt and vaccination interval are tightly linked variables

It has been previously demonstrated that longer intervals between exposures, whether through natural infection or vaccination, may result in enhanced potency and breadth of neutralizing antibody responses (*19–21*). Compared to bivalent recipients, the non-recipient group had a greater interval between the XBB.1.5 monovalent vaccine and prior vaccination (mean of 361.2 ± 48.3 days versus mean of 706.9 ± 58.9 days). It is therefore of interest to evaluate the contribution of these longer intervals to the increased boosting observed in bivalent non-recipients. However, despite being statistically significant, representative multiple predictors linear regression modeling on EG.5.1 titer fold-change revealed that bivalent receipt and vaccine interval were highly colinear variables within our cohort as evident in variance inflation factors >10 and R^2^ > 0.9 between variables (**Supplemental Figure 3**). In other words, those who opted to not receive the bivalent vaccine reliably had longer rest intervals between vaccines. This is an expected problem due to the inherently intertwined nature of these variables in real-world vaccinated communities and indicates that regression modeling within this cohort cannot be used fully isolate the independent effect of vaccine intervals on antibody boosting. However, it is important to emphasize that ancestral titers are independently “back-boosted” within each participant group, providing evidence that immune imprinting persists in the context of XBB.1.5 monovalent vaccination regardless of the differences, including vaccine intervals, between the two groups and is likely remnant of original ancestral monovalent vaccines (Figure 1B, 1C, 1D, 1E).

## DISCUSSION

We examined variant-specific humoral responses to XBB.1.5 monovalent vaccination in individuals who did and did not receive prior boosting with the bivalent Wuhan-Hu-1/BA.4-5 vaccine. In each group, XBB.1.5 vaccination resulted in significant boosting of humoral immunity as measured by RBD-specific IgG, IgA, and total IgG/A/M isotypes and neutralizing titers against XBB.1.5, EG.5.1, and JN.1. Importantly, both groups exhibited “back-boosting” of neutralizing titers against the ancestral WA1 strain and isotype titers binding the ancestral spike RBD, which is highly characteristic of persistent immune imprinting (*8*). While EG.5.1 was neutralized by vaccinated sera to the same degree as XBB.1.5, JN.1 demonstrated marked escape. These data indicate that while the XBB.1.5 vaccine is highly immunogenic, additional updates will be required to counter continued viral evolution. Indeed, the JN.1 lineage gave rise to currently circulating variants, including KP.2, KP.3, LP.8.1, and the dominant XEC strains (*22*). At post-XBB.1.5 vaccination, neutralizing titers against KP.2 appear to fall 3.1-fold compared to JN.1, with KP.3.1.1 demonstrating similar evasion of neutralization (*2*, *23*). Thus, vaccines have been formulated to target this emerging lineage of variants and are being rolled out at present; however, our findings suggest that imprinting and vaccination intervals will be important considerations for these and future updates. In particular, the effectiveness of JN.1 formulations should be evaluated in terms of immune imprinting from prior vaccines, including the 2023 XBB.1.5 monovalent vaccine assessed herein.

Beyond “back-boosting,” we demonstrate that immune imprinting also affects responses to the updated XBB.1.5 vaccine by direct comparisons to a group of participants missing the prior bivalent vaccine. Experiencing less imprinting upon XBB.1.5 vaccination, these bivalent non-recipients had greater boosting of neutralizing antibodies with trends toward higher absolute titers and comparable cross-neutralization. Within the bivalent recipient group, those who had one less ancestral monovalent vaccine also appear to exhibit greater boosting of XBB.1.5 and EG.5.1 neutralization after XBB.1.5 vaccination. This is in line with previous reports that repetitive vaccination with the ancestral spike protein causes profound immune imprinting (*24*). Indeed, despite the exclusion of ancestral Wuhan-Hu-1 spike protein in the XBB.1.5 monovalent vaccine, XBB.1.5 vaccination largely results in recall of ancestral spike-reactive memory B cells instead of generating novel responses (*25*). Fortunately, additional Omicron exposures through natural infection or vaccination may break immune imprinting by ancestral spike protein (*26*). Therefore, vaccines that exclude historic variant spike proteins will be preferable to minimize imprinting, though multivalent formulations may still be effective if they combine circulating variants (*27*). Encouragingly, responses against JN.1 were similar whether bivalent recipients received 3 or 4 ancestral vaccinations, suggesting that the grip of ancestral imprinting on vaccine immunogenicity against this and more contemporary lineages may be weakening.

We further demonstrated that vaccine intervals and bivalent receipt were highly colinear variables in our cohort and likely in the vaccinated community, prohibiting the isolation of each factor’s independent contribution to enhanced bivalent non-recipient boosting. However, longer exposure intervals have been extensively shown to enhance humoral immunity against SARS-CoV-2 variants and likely contributes to enhanced non-recipient boosting (*19*, *20*). Our previous study similarly demonstrated this interval effect, but with participants that had up to 400 days between exposures (*21*). This suggests that a fruitful strategy for COVID-19 vaccines will be to optimize recommended intervals while minimizing disease risk from waning immunity. Of particular interest will be comprehensive studies characterizing efficacy and risk when extending the current trend of annual recommendations to longer, perhaps biennial intervals.

While we observed that bivalent non-recipients experience greater potency and boosting of neutralizing antibodies, it is important to emphasize that this group had lower pre-vaccination titers against variants, cross-neutralization, and neutralizing potency due to waning immunity and lack of Omicron boosting by the BA.4/5 spike variant. Though both groups had similar infection histories, these diminished responses underline a possible increase in infection susceptibility in bivalent non-recipients before XBB.1.5 vaccination. Indeed, prior to being excluded from the final cohort based on anti-nucleocapsid positivity, 2/12 (16.7%) participants in the initial bivalent non-recipient group compared to 3/37 (8.1%) in the initial recipient group were infected in between serum collection timepoints before the XBB.1.5 vaccination could sufficiently boost adaptive immunity. These trends highlight the risk we must avoid when optimizing the current vaccine strategy. Importantly, current annual vaccines are highly effective at reducing disease burden with estimates of XBB.1.5 vaccine effectiveness against COVID-19 hospitalization ranging from 43% to 76.1% (*28*, *29*). Collectively, the foregoing observations support public health recommendations for remaining current with updated vaccinations to protect against disease caused by novel, emerging variants.

The primary limitation of this study is a small sample size for the bivalent non-recipient group, which is comprised of relatively rare individuals missing only the 2022 bivalent vaccine and opting for the 2023 formulation. However, our assessment of immune imprinting on XBB.1.5 vaccine immunogenicity is corroborated by orthogonal studies with larger cohorts (*5*, *25*). Furthermore, vaccine interval and bivalent receipt are highly colinear within our cohort, limiting interval-independent observations of imprinting to ancestral “back-boosting” meanwhile extended intervals may contribute to increased non-recipient boosting. The isolation of imprinting influence on this particular phenotype will thus require granular cohorts with more overlapping vaccine intervals. We evaluated only antibody responses, and other immune facets such as T cell activity, memory B cell response, and non-neutralizing Fc effector functions will need to be assessed in the context of immune imprinting. Despite our study limitations, we demonstrate immune imprinting through “back-boosting” of ancestral titers and, most importantly, direct comparisons of groups with fewer imprinting vaccines. Of greatest need will be studies connecting these diminished immune responses to clinical outcomes while controlling for selection bias.

## MATERIALS AND METHODS

### Study Design

The purpose of this study was to compare humoral immune responses to the XBB.1.5 monovalent vaccine in individuals who received all United States Center for Disease Control and Prevention (CDC)-recommended COVID-19 vaccines since the beginning of the pandemic versus those who were missing just the bivalent Wuhan-Hu-1/BA.4-5 vaccine in 2022. Serum samples were collected from participants before and after XBB.1.5 vaccination, which were assessed using ELISAs and FRNTs. Bivalent non-recipients were selected for inclusion based on having received all prior CDC-recommended doses except for the bivalent vaccine. Bivalent recipient controls were selected on the basis of prior vaccine history, sex, age, and days since receiving the bivalent vaccine in order to closely match demographics in the non-recipient group. Number of prior infections, suspected and confirmed, were similar across both groups. All participant samples were tested for anti-nucleocapsid antibodies by ELISA to screen for recent infection and excluded from analysis if deemed positive.

### Cohort Selection and Serum Collection

Healthcare workers at Oregon Health and Science University (OHSU) were recruited and enrolled in the study. Written informed consent was obtained at the time of enrollment. Study approval was obtained from the OHSU institutional review board (IRB). This study was carried out in compliance with all relevant ethical regulations as outlined by the IRB. Bivalent recipients were fully vaccinated, defined as having received 3 or 4 doses of ancestral Wuhan-Hu-1 vaccine and 1 dose of bivalent Wuhan-Hu-1/BA.4-5 vaccine. Bivalent non-recipients were defined as missing the bivalent vaccine but otherwise fully vaccinated. At the time of enrollment, each participant contributed 4-6mL of whole blood, which was then centrifuged at 1000 x g, subjected to heat inactivation at 56°C for 30 minutes, and subsequently stored at -20°C to serve as a baseline (pre-boost) serum sample. Roughly three weeks following the administration of the XBB.1.5-adapted monovalent vaccine (Moderna Therapeutics), additional whole blood samples were collected and processed in the same manner to obtain post-boost serum samples.

### Cell Culture

Vero E6 monkey kidney epithelial cells (ATCC, CRL-1586) were maintained in tissue culture-treated plates in Dulbecco’s modified Eagle’s medium (DMEM), 10% fetal bovine serum (FBS), 1% nonessential amino acids (NEAAs), and 1% penicillin-streptomycin (PS) at 37C, 5% CO2, 100% relative humidity.

### SARS-CoV-2 Growth and Titration

SARS-CoV-2 clinical isolates were obtained from BEI Resources: WA1, USAWA1/2020 (NR-52281); XBB.1.5, USA/MD-HP40900/2022 (NR-59104); EG.5.1, USA/MD-HP47946/2023 (NR-59503); and JN.1, USA/New York/PV96109/2023 (NR-59693). To generate Passage 1 viruses, ∼70% confluent Vero E6 cells were inoculated and cultured until cytopathic effect appeared—72 hours for WA1 and 96 hours for XBB.1.5, EG.5.1, and JN.1. The culture supernatants were collected, centrifuged at 1000 x g for 10 minutes, and then stored in aliquots at -80°C. Passage 2 viruses were produced in a similar manner using Passage 1 viruses for inoculation.

Virus titers were established via a focus-forming assay with 10-fold dilutions in dilution medium (2% FBS, 1% NEAA, and 1% P/S in Opti-MEM). Confluent Vero E6 cells in 96-well plates were incubated with virus dilutions for one hour at 37°C in 5% CO2. Overlay medium (1% methylcellulose in dilution medium) was then added, and plates were incubated for 24 hours for WA1 and 48 hours for the other strains. Following this, the overlay medium was removed, and cells were fixed using 4% formaldehyde in PBS for one hour at RT. To stain infection foci, cells were permeabilized in perm buffer (0.1% BSA and 0.1% saponin in PBS) for 30 minutes at RT and then incubated with polyclonal anti-SARS-CoV-2 alpaca serum diluted at 1:5000 for WA1 and 1:500 for XBB.1.5, EG.5.1, and JN.1 overnight at 4°C. After washing three times with wash buffer (0.01% Tween-20 in PBS) cells were treated with HRP-conjugated anti-alpaca IgG (Novus #NB7242) diluted at 1:20,000 for WA1 and 1:10,000 for the other strains for two hours at RT. Following another three washes, 50µL of KPL TrueBlue substrate (Seracare #5510-0030) was added per well. After 20 minutes of incubation at room temperature, plates were imaged using a CTL Immunospot Analyzer. Foci were then counted with the Viridot package for R 4.3.1 (*30*).

### ELISAs

ELISAs for each Ig isotype were conducted in biological duplicates in 96-well plates (Maxisorp #423501). Each plate was coated overnight at 4°C with 100µL per well of purified wild-type SARS-CoV-2 RBD at 1µg/mL in PBS, with gentle rocking. The plates were washed three times with wash buffer (0.05% Tween-20 in PBS), then incubated with 150µL per well of blocking buffer (5% nonfat dry milk powder and 0.05% Tween-20 in PBS) for 1 hour at RT, rocking. Participant serum samples were diluted in blocking buffer to create five 4-fold serial dilutions (from 1:25 to 1:6,400). Plates were incubated with serum dilutions for one hour at RT, rocking, followed by three washes. HRP-conjugated secondary antibodies against human IgG (BD Biosciences #555788), IgA (BioLegend #411002), and IgM (Bethyl Laboratories #A80-100P) were used at a 1:3,000 dilution in blocking buffer, and 100µL was added per well. For total antibody detection, HRP-conjugated goat anti-human IgG/A/M (Invitrogen, #A18847) was used at a 1:10,000 dilution. After 1 hour at RT, rocking, and three washes, o-phenylenediamine dihydrochloride (ThermoFisher Scientific, #34005) was added as per the manufacturer’s instructions. The reaction was halted after 25 minutes with an equal volume of 1M HCl, and the optical density (OD) was measured at 492nm on a CLARIOstar plate reader. OD492 values were normalized by subtracting the mean of the negative control wells. The protocol for detecting anti-nucleocapsid antibodies was identical, except the plates were coated with 1µg/mL nucleocapsid protein (SinoBio #40588-V08B) and detected using HRP goat anti-human IgG/A/M. Serum was deemed positive for anti-nucleocapsid antibodies if the calculated EC50 exceeded the lower limit of detection of 25.

### SARS-CoV-2 FRNT

Focus reduction neutralization tests (FRNT) were performed as previously described (*31*). Briefly, participant serum samples were prepared in duplicate, with five 10-fold serial dilutions for WA1 (1:10 to 1:100,000) and five 5-fold serial dilutions for XBB.1.5, EG.5.1, and JN.1 (from 1:10 to 1:6250). These dilutions were mixed with equal volume of 2X concentrated virus and incubated for 1 hour at RT. The mixtures were then added to Vero E6 cells in 96-well plates and incubated at 37°C with 5% CO2 for 1 hour, followed by a further incubation with overlay media for 24 hours for WA1 and 48 hours for XBB.1.5, EG.5.1, and JN.1. Cells were then fixed with 4% formaldehyde in PBS for one hour at room temperature. All subsequent staining, developing, imaging, and counting followed methods used in prior titration experiments. For FRNT, approximately 30 focus-forming units (FFU) of virus per well were used, and for each set of five test wells, a sixth control well without serum was included. Percent neutralization for each test well was calculated by comparing the focus count to the average in the control wells.

### ELISA EC50 and FRNT50 Calculations

ELISA OD492 readings and FRNT percent neutralization metrics were analyzed using Python (v3.7.6) with the data analysis libraries numpy (v1.18.1), scipy (v1.4.1), Matplotlib (v3.1.3), and pandas (v1.0.1) to compute ELISA EC50 and FRNT50 values. Data from replicates were combined and fit using a three-parameter logistic model. Replicate curves for both ELISA EC50 and FRNT50 were generated separately for quality control. If the final ELISA EC50 values were below the quantification limit of 25, they were adjusted to 24; values exceeding the upper limit of 6,400 were adjusted to 6,401. Similarly, final FRNT50 values below the quantification threshold of 20 were adjusted to 19, and values surpassing the upper limits of 200,000 for WA1 and 12,500 for XBB.1.5, EG.5.1, and JN.1 were set to 200,001 and 12,501, respectively.

### Antigenic Cartography

FRNT50 neutralizing titer values were processed and analyzed using published methods for antigenic cartography (*32*). Using the Racmacs package (v1.2.9, https://acorg.github.io/Racmacs/) for R (v.4.3.1), antigenic distances between serum samples and SARS-CoV-2 strains were calculated and visualized. For 2-dimensional mapping of serum and variants for each study group (bivalent recipients or non-recipients) and at each timepoint (pre- and post-vaccination), optimization was set to 2000 steps and “minimum column basis” was set to “none.” Manual seeds were set for each map to orient WA1 to the left side of each plot. Map distances between WA1 and each variant were extracted with the dist(ptCoords()) function.

### Statistical Analysis

For repeated measures comparisons involving antibody titers (EC50), a restricted effect maximum likelihood (REML) model with Holm-Sidak multiple comparisons correction were performed on the log-transformed data. For similar comparisons involving neutralizing antibody titers, a repeated measures one-way ANOVA with Holm-Sidak multiple comparisons correction were performed on log-transformed FRNT50 data. All other multi-group statistics were assessed by standard one-way ANOVA with follow-up “tests for linear trend” (a.k.a. “test for linear contrast,” “post-test for trend”), which assesses systematic increases or decreases in column means in left-to-right column order. Single pairwise comparisons of absolute titers or fold changes between groups were the result of two-tailed, unpaired t tests. Multiple linear regression modeling was performed on log-transformed fold-change (post/pre-vaccination titer) data with bivalent receipt and vaccine interval as explanatory variables. Outliers were identified and removed by Robust Regression and Outlier Removal (ROUT) with Q=0.1%. P-values less than 0.05 were considered statistically significant. All statistical analyses were performed in GraphPad Prism v10.

## Data Availability

The authors declare that the data associated with this study are provided within the paper or in public repositories. FRTN50 and EC50 values will be made publicly available on Zenodo upon publication.

## List of Supplemental Materials

**Supplemental Fig. 1**. Pre- and post-XBB.1.5 vaccination neutralizing titers against SARS-CoV-2 variants stratified by ancestral vaccine history.

**Supplemental Fig. 2**. Pre- and post-XBB.1.5 vaccination antibody isotype titers against ancestral spike RBD stratified by ancestral vaccine history.

**Supplemental Fig. 3**. Bivalent receipt and vaccine interval are highly colinear variables in representative EG.5.1 regression modeling.

### Code availability

The FRNT50/EC50 calculation code used in this study is available on Github at DOI: 10.5281/zenodo.5158655.

## Acknowledgements

The authors thank the many participants in this study for their generous contribution. We gratefully acknowledge the efforts of the entire OHSU COVID-19 serology study team. We also express gratitude to Lauren Rodda for helpful discussions.

## Author contributions

Conceptualization: XW, MC, FT

Methodology: XW, TB, MT, MW, AH, MC, FT

Investigation: XW, TB, MT, MW, AH

Visualization: XW

Funding acquisition: MC, FT

Project administration: XW, MT, MW, AH

Supervision: MC, FT

Writing – original draft: XW

Writing – review & editing: XW, TB, MT, MW, AH, MC, FT

### Materials and correspondence

For any materials requests or correspondence, please contact Fikadu G. Tafesse.

## References

1. E. Harris, WHO Declares End of COVID-19 Global Health Emergency. JAMA 329, 1817 (2023).

2. Y. Kaku, K. Uriu, Y. Kosugi, K. Okumura, D. Yamasoba, Y. Uwamino, J. Kuramochi, K. Sadamasu, K. Yoshimura, H. Asakura, M. Nagashima, T. G. to P. J. (G2P-J. Consortium, J. Ito, K. Sato, Virological characteristics of the SARS-CoV-2 KP.2 variant. bioRxiv [Preprint] (2024). 10.1101/2024.04.24.590786.

3. M. V. Stankov, M. Hoffmann, R. G. Jauregui, A. Cossmann, G. M. Ramos, T. Graalmann, E. J. Winter, M. Friedrichsen, I. Ravens, T. Ilievska, J. Ristenpart, A. Schimrock, S. Willenzon, G. Ahrenstorf, T. Witte, R. Förster, A. Kempf, S. Pöhlmann, S. I. Hammerschmidt, A. Dopfer-Jablonka, G. M. N. Behrens, Humoral and cellular immune responses following BNT162b2 XBB.1.5 vaccination. Lancet Infect. Dis. 24, e1–e3 (2024).

4. N. Patel, J. F. Trost, M. Guebre-Xabier, H. Zhou, J. Norton, D. Jiang, Z. Cai, M. Zhu, A. M. Marchese, A. M. Greene, R. M. Mallory, R. Kalkeri, F. Dubovsky, G. Smith, XBB.1.5 spike protein COVID-19 vaccine induces broadly neutralizing and cellular immune responses against EG.5.1 and emerging XBB variants. Sci. Rep. 13, 19176 (2023).

5. Q. Wang, Y. Guo, A. Bowen, I. A. Mellis, R. Valdez, C. Gherasim, A. Gordon, L. Liu, D. D. Ho, XBB.1.5 monovalent mRNA vaccine booster elicits robust neutralizing antibodies against XBB subvariants and JN.1. Cell Host Microbe 32, 315–321.e3 (2024).

6. X. H. Nguyenla, T. A. Bates, M. Trank-Greene, M. Wahedi, F. G. Tafesse, M. Curlin, Humoral Immunity Elicited by the XBB.1.5 Monovalent COVID-19 Vaccine. medRxiv [Preprint] (2024). 10.1101/2024.03.25.24304857.

7. C. for B. E. and Research, Updated COVID-19 Vaccines for Use in the United States Beginning in Fall 2024. FDA (2024).

8. M. Koutsakos, A. H. Ellebedy, Immunological imprinting: Understanding COVID-19. Immunity 56, 909–913 (2023).

9. T. Aydillo, A. Rombauts, D. Stadlbauer, S. Aslam, G. Abelenda-Alonso, A. Escalera, F. Amanat, K. Jiang, F. Krammer, J. Carratala, A. García-Sastre, Immunological imprinting of the antibody response in COVID-19 patients. Nat. Commun. 12, 3781 (2021).

10. C. J. Reynolds, C. Pade, J. M. Gibbons, A. D. Otter, K.-M. Lin, D. Muñoz Sandoval, F. P. Pieper, D. K. Butler, S. Liu, G. Joy, N. Forooghi, T. A. Treibel, C. Manisty, J. C. Moon, COVIDsortium Investigators, COVIDsortium Immune Correlates Network, A. Semper, T. Brooks, Á. McKnight, D. M. Altmann, R. J. Boyton, Immune boosting by B.1.1.529 (Omicron) depends on previous SARS-CoV-2 exposure. Science 377, eabq1841 (2022).

11. K. Röltgen, S. C. A. Nielsen, O. Silva, S. F. Younes, M. Zaslavsky, C. Costales, F. Yang, O. F. Wirz, D. Solis, R. A. Hoh, A. Wang, P. S. Arunachalam, D. Colburg, S. Zhao, E. Haraguchi, A. S. Lee, M. M. Shah, M. Manohar, I. Chang, F. Gao, V. Mallajosyula, C. Li, J. Liu, M. J. Shoura, S. B. Sindher, E. Parsons, N. J. Dashdorj, N. D. Dashdorj, R. Monroe, G. E. Serrano, T. G. Beach, R. S. Chinthrajah, G. W. Charville, J. L. Wilbur, J. N. Wohlstadter, M. M. Davis, B. Pulendran, M. L. Troxell, G. B. Sigal, Y. Natkunam, B. A. Pinsky, K. C. Nadeau, S. D. Boyd, Immune imprinting, breadth of variant recognition, and germinal center response in human SARS-CoV-2 infection and vaccination. Cell 185, 1025–1040.e14 (2022).

12. Collier Ai-ris Y., Miller Jessica, Hachmann Nicole P., McMahan Katherine, Liu Jinyan, Bondzie Esther A., Gallup Lydia, Rowe Marjorie, Schonberg Eleanor, Thai Siline, Barrett Julia, Borducchi Erica N., Bouffard Emily, Jacob-Dolan Catherine, Mazurek Camille R., Mutoni Audrey, Powers Olivia, Sciacca Michaela, Surve Nehalee, VanWyk Haley, Wu Cindy, Barouch Dan H., Immunogenicity of BA.5 Bivalent mRNA Vaccine Boosters. N. Engl. J. Med. 388, 565–567 (2023).

13. Wang Qian, Bowen Anthony, Valdez Riccardo, Gherasim Carmen, Gordon Aubree, Liu Lihong, Ho David D., Antibody Response to Omicron BA.4–BA.5 Bivalent Booster. N. Engl. J. Med. 388, 567–569 (2023).

14. Q. Wang, A. Bowen, A. R. Tam, R. Valdez, E. Stoneman, I. A. Mellis, A. Gordon, L. Liu, D. D. Ho, SARS-CoV-2 neutralising antibodies after bivalent versus monovalent booster. Lancet Infect. Dis. 23, 527–528 (2023).

15. T. Cerqueira-Silva, V. S. Boaventura, M. Barral-Netto, Effectiveness of monovalent and bivalent COVID-19 vaccines. Lancet Infect. Dis. 23, 1208–1209 (2023).

16. H. Chemaitelly, H. H. Ayoub, P. Tang, P. V. Coyle, H. M. Yassine, A. A. Al Thani, H. A. Al-Khatib, M. R. Hasan, Z. Al-Kanaani, E. Al-Kuwari, A. Jeremijenko, A. H. Kaleeckal, A. N. Latif, R. M. Shaik, H. F. Abdul-Rahim, G. K. Nasrallah, M. G. Al-Kuwari, A. A. Butt, H. E. Al-Romaihi, M. H. Al-Thani, A. Al-Khal, R. Bertollini, L. J. Abu-Raddad, History of primary-series and booster vaccination and protection against Omicron reinfection. Sci. Adv. 9, eadh0761 (2023).

17. C. H. Hansen, Evidence of immune imprinting or the effect of selection bias? Sci. Adv. 9, eadk5668 (2023).

18. D. Planas, I. Staropoli, V. Michel, F. Lemoine, F. Donati, M. Prot, F. Porrot, F. Guivel-Benhassine, B. Jeyarajah, A. Brisebarre, O. Dehan, L. Avon, W. H. Bolland, M. Hubert, J. Buchrieser, T. Vanhoucke, P. Rosenbaum, D. Veyer, H. Péré, B. Lina, S. Trouillet-Assant, L. Hocqueloux, T. Prazuck, E. Simon-Loriere, O. Schwartz, Distinct evolution of SARS-CoV-2 Omicron XBB and BA.2.86/JN.1 lineages combining increased fitness and antibody evasion. Nat. Commun. 15, 2254 (2024).

19. B. Grunau, D. M. Goldfarb, M. Asamoah-Boaheng, L. Golding, T. L. Kirkham, P. A. Demers, P. M. Lavoie, Immunogenicity of Extended mRNA SARS-CoV-2 Vaccine Dosing Intervals. JAMA 327, 279–281 (2022).

20. V. G. Hall, V. H. Ferreira, H. Wood, M. Ierullo, B. Majchrzak-Kita, K. Manguiat, A. Robinson, V. Kulasingam, A. Humar, D. Kumar, Delayed-interval BNT162b2 mRNA COVID-19 vaccination enhances humoral immunity and induces robust T cell responses. Nat. Immunol. 23, 380–385 (2022).

21. T. A. Bates, H. C. Leier, S. K. McBride, D. Schoen, Z. L. Lyski, D. X. Lee, W. B. Messer, M. E. Curlin, F. G. Tafesse, An extended interval between vaccination and infection enhances hybrid immunity against SARS-CoV-2 variants. JCI Insight 8 (2023).

22. COVID-19 variants | WHO COVID-19 dashboard, datadot. https://data.who.int/dashboards/covid19/variants.

23. Y. Kaku, K. Uriu, K. Okumura, J. Ito, K. Sato, Virological characteristics of the SARS-CoV-2 KP.3.1.1 variant. Lancet Infect. Dis. 0 (2024).

24. Q. Wang, Y. Guo, A. R. Tam, R. Valdez, A. Gordon, L. Liu, D. D. Ho, Deep immunological imprinting due to the ancestral spike in the current bivalent COVID-19 vaccine. Cell Rep. Med. 4, 101258 (2023).

25. M. A. Tortorici, A. Addetia, A. J. Seo, J. Brown, K. Sprouse, J. Logue, E. Clark, N. Franko, H. Chu, D. Veesler, Persistent immune imprinting occurs after vaccination with the COVID-19 XBB.1.5 mRNA booster in humans. Immunity 57, 904–911.e4 (2024).

26. A. Yisimayi, W. Song, J. Wang, F. Jian, Y. Yu, X. Chen, Y. Xu, S. Yang, X. Niu, T. Xiao, J. Wang, L. Zhao, H. Sun, R. An, N. Zhang, Y. Wang, P. Wang, L. Yu, Z. Lv, Q. Gu, F. Shao, R. Jin, Z. Shen, X. S. Xie, Y. Wang, Y. Cao, Repeated Omicron exposures override ancestral SARS-CoV-2 immune imprinting. Nature 625, 148–156 (2024).

27. S. Chang, K.-S. Shin, B. Park, S. Park, J. Shin, H. Park, I. K. Jung, J. H. Kim, S. E. Bae, J.-O. Kim, S. H. Baek, G. Kim, J. J. Hong, H. Seo, E. Volz, C.-Y. Kang, Strategy to develop broadly effective multivalent COVID-19 vaccines against emerging variants based on Ad5/35 platform. Proc. Natl. Acad. Sci. 121, e2313681121 (2024).

28. C. H. Hansen, I. R. Moustsen-Helms, M. Rasmussen, B. Søborg, H. Ullum, P. Valentiner-Branth, Short-term effectiveness of the XBB.1.5 updated COVID-19 vaccine against hospitalisation in Denmark: a national cohort study. Lancet Infect. Dis. 0 (2024).

29. J. DeCuir, Interim Effectiveness of Updated 2023–2024 (Monovalent XBB.1.5) COVID-19 Vaccines Against COVID-19–Associated Emergency Department and Urgent Care Encounters and Hospitalization Among Immunocompetent Adults Aged ≥18 Years — VISION and IVY Networks, September 2023–January 2024. MMWR Morb. Mortal. Wkly. Rep. 73 (2024).

30. L. C. Katzelnick, A. C. Escoto, B. D. McElvany, C. Chávez, H. Salje, W. Luo, I. Rodriguez-Barraquer, R. Jarman, A. P. Durbin, S. A. Diehl, D. J. Smith, S. S. Whitehead, D. A. T. Cummings, Viridot: An automated virus plaque (immunofocus) counter for the measurement of serological neutralizing responses with application to dengue virus. PLoS Negl. Trop. Dis. 12, e0006862 (2018).

31. T. A. Bates, H. C. Leier, Z. L. Lyski, S. K. McBride, F. J. Coulter, J. B. Weinstein, J. R. Goodman, Z. Lu, S. A. R. Siegel, P. Sullivan, M. Strnad, A. E. Brunton, D. X. Lee, A. C. Adey, B. N. Bimber, B. J. O’Roak, M. E. Curlin, W. B. Messer, F. G. Tafesse, Neutralization of SARS-CoV-2 variants by convalescent and BNT162b2 vaccinated serum. Nat. Commun. 12, 5135 (2021).

32. D. J. Smith, A. S. Lapedes, J. C. de Jong, T. M. Bestebroer, G. F. Rimmelzwaan, A. D. M. E. Osterhaus, R. A. M. Fouchier, Mapping the Antigenic and Genetic Evolution of Influenza Virus. Science 305, 371–376 (2004).

