## Supplemental Data for "Immune imprinting and vaccination interval underly XBB.1.5 monovalent vaccine immunogenicity"

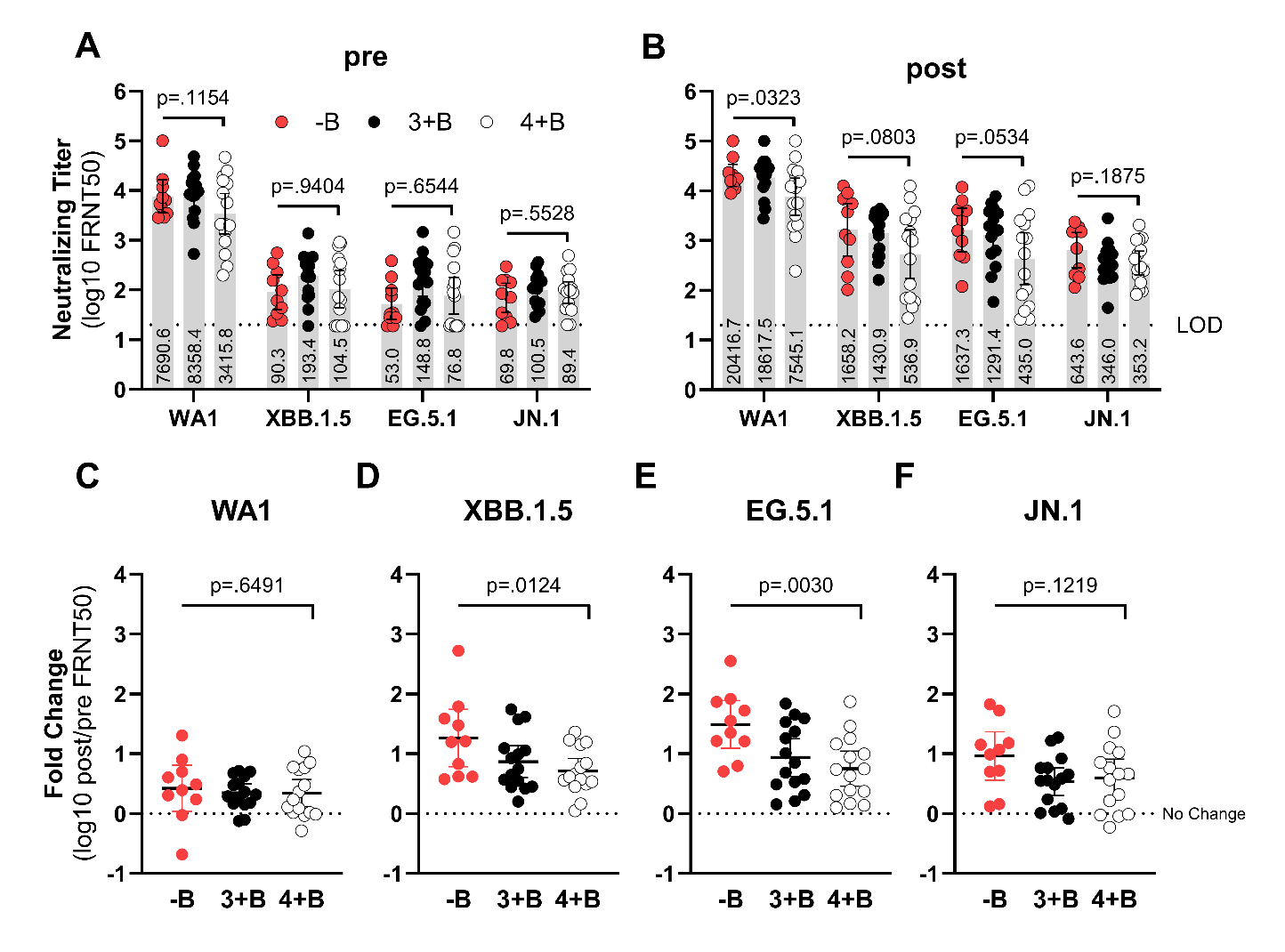

**Supplemental Fig. 1. Pre- and post-XBB.1.5 vaccination neutralizing titers against SARS-CoV-2 variants stratified by ancestral vaccine history.** Bivalent recipients were divided based upon receiving 3 (3+B) or 4 (4+B) ancestral vaccine doses prior to their bivalent vaccine; bivalent non-recipients (-B). Live SARS-CoV-2 neutralization by serum antibodies was assessed by FRNT and reported as FRNT50s at pre- (**A**) or post-XBB.1.5 vaccination (**B**). The dotted lines indicate assay lower limits of detection. Numerical values for geometric mean titers (GMTs) are displayed. Fold changes were calculated by dividing the post-XBB.1.5 vaccination titer by pre-vaccination titer for each participant against WA1 (**C**), XBB.1.5 (**D**), EG.5.1 (**E**), and JN.1 (**F**). For all plots, error bars are geometric means with 95% confidence intervals. Reported p-values are the result of one-way ANOVA with column tests for linear trend.

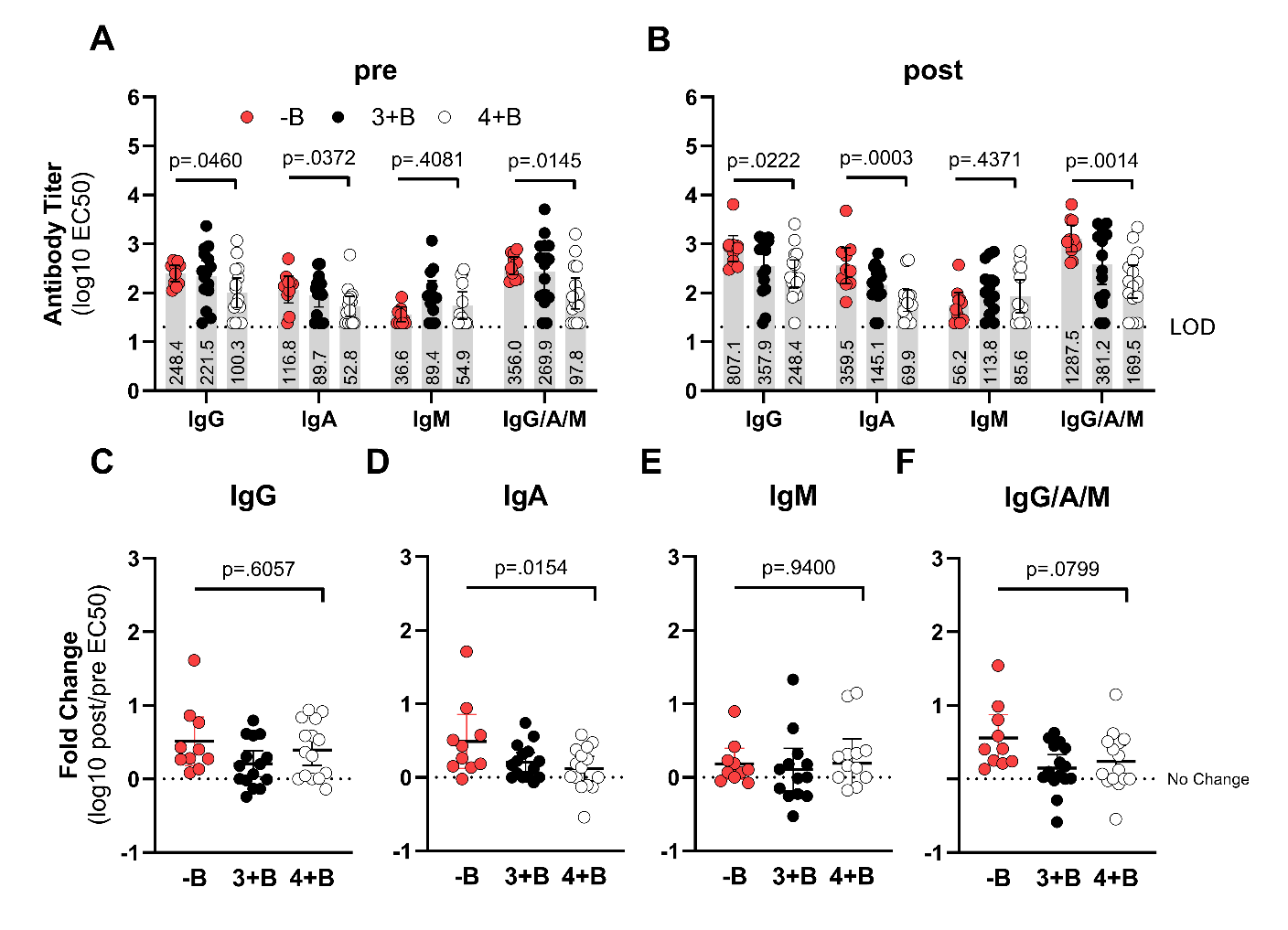

**Supplemental Fig. 2. Pre- and post-XBB.1.5 vaccination antibody isotype titers against ancestral spike RBD stratified by ancestral vaccine history.** Bivalent recipients were divided based upon receiving 3 (3+B) or 4 (4+B) ancestral vaccine doses prior to their bivalent vaccine; bivalent non-recipients (-B). Serum antibody isotype titers against ancestral spike RBD were determined by ELISA and reported as EC50 at pre- (**A**) or post-XBB.1.5 vaccination (**B**). The dotted lines indicate assay lower limits of detection. Numerical values for geometric mean titers (GMTs) are displayed. Fold changes were calculated by dividing the post-XBB.1.5 vaccination titer by pre-vaccination titer for each participant for IgG (**C**), IgA (**D**), IgM (**E**), and total IgG/A/M (**F**). For all plots, error bars are geometric means with 95% confidence intervals. Reported p-values are the result of one-way ANOVA with column tests for linear trend.

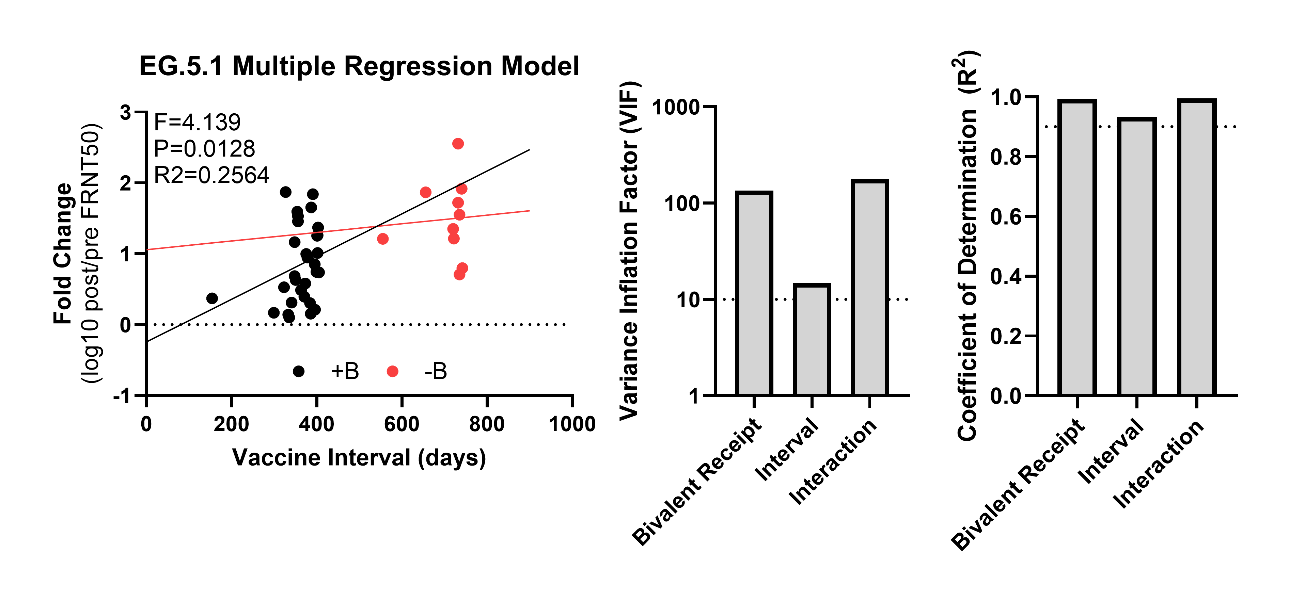

**Supplemental Fig. 3.** **Bivalent receipt and vaccine interval are highly colinear variables in representative EG.5.1 regression modeling.** Multiple predictors linear regression modeling was performed within Graphpad Prism with fold-change neutralizing titers against EG.5.1 as the dependent variable and with bivalent vaccination status and interval between XBB.1.5 and prior vaccination as explanatory variables. The overall regression model exhibited a statistically significant weak, positive association between antibody fold-change and combined bivalent receipt, vaccine interval, and interaction variables. Regression lines for each group were produced by fixing the bivalent receipt term then interpolating fold-changes. Diagnosis of collinearity between variables was performed through calculation of variance inflation factors (VIF) for each variable and coefficients of determination (R^2^) between explanatory variables; dotted lines for VIF>10 and R^2^>0.9 represent cutoff for severe collinearity.
